# Lives saved and hospitalizations averted by COVID-19 vaccination in New York City

**DOI:** 10.1101/2021.07.14.21260481

**Authors:** Affan Shoukat, Thomas N. Vilches, Seyed M. Moghadas, Pratha Sah, Eric C. Schneider, Jaimie Shaff, Jane Zucker, Celia Quinn, Dave A. Chokshi, Alison P. Galvani

## Abstract

Despite the emergence of highly transmissible variants, the number of cases in NYC has fallen from over 5,500 average daily cases in January, 2020 to less than 350 average daily cases in July, 2021. The impact of vaccination in saving lives and averting hospitalizations in NYC has not been formally investigated yet. We used an age-stratified agent-based model calibrated to COVID-19 transmission and vaccination in NYC to evaluate the impact of the vaccination campaign in suppressing the COVID-19 burden. We found that the vaccination campaign has prevented over 250,000 COVID-19 cases, 44,000 hospitalizations and 8,300 deaths from COVID-19 infection since the start of vaccination through July 1, 2021. Notably, the swift vaccine rollout suppressed another wave of COVID-19 that would have led to sustained increase in cases, hospitalizations and deaths during spring triggered by highly transmissible variants. As the Delta variant sweeps across the city, the findings of this study underscore the urgent need to accelerate vaccination and close the vaccine coverage gaps across the city.

## Background

New York City (NYC) was one of the hardest-hit cities in the US during the initial phase of the COVID-19 pandemic. Following multiple early introductions of the virus from different regions globally, the metropolis faced an arduous challenge in controlling rapid community spread. Soon after its first COVID-19 case was identified on 29 February 2020,^1^ the number of cases detected rose precipitously. Within two months the city had turned into the world’s epicenter of the COVID-19 pandemic, reporting over 100,000 cases, accounting for 21% of cases in the US and 7% worldwide at that time.

Vaccinations in NYC began in December 2020^2^ and as of July 11, 2021, NYC has distributed 9.6 million doses of vaccines, achieving vaccination of 68.9% of its 6.6 million adult residents with at least one dose.^2^ Compared with the national average of 59% full vaccinated adults, NYC has fully vaccinated 64% of adults.^2^ At the same time, cases in NYC have declined from more than 5,500 average daily cases in January, 2020 to less than 350 average daily cases in July, 2021 despite emergence of highly transmissible variants.^3^ The impact of vaccination in saving lives and averting hospitalizations in NYC has not been formally investigated yet.

## Methods

To estimate the impact of the vaccination program as was implemented in NYC, we extended our previously published an age-stratified agent-based model of COVID-19 disease dynamics, ^4^ including transmission characteristics of Alpha (B.1.1.7), Gamma (P.1), and Delta (B.1.617.2) variants as identified in NYC. Model parameters included city-specific population demographics, age-specific risks of severe outcomes and hospitalization, a contact network accounting for mobility patterns during the pandemic, and the natural history of COVID-19 disease. An age-specific, two-dose vaccination strategy was implemented in which vaccine administration and allocation followed the daily rate of vaccination in NYC stratified by age groups starting on December 14, 2020.^5^ We considered vaccine efficacies against transmission, symptomatic infection, and severe disease, with parameter values derived from published estimates (Appendix, Table S3). The model was calibrated to reported incidence in NYC from October 1, 2020 to July 1, 2021, accounting for the time of identification of the Alpha (January 3, 2021), Gamma (March 20, 2021), and Delta (Feb 20, 2021) variants as well as current vaccination dynamics. With the parameters calibrated to vaccination rollout in NYC, we then simulated the model under the counterfactual scenario of no vaccination and estimated the number of cases, hospitalizations and deaths have been averted due to rapid vaccination of the population.

## Results

Without vaccination, we estimated that an average of 851,235 infections, 86,858 hospitalizations, and 17,265 deaths would have occurred in NYC between December 12, 2020 to July 1, 2021. The COVID-19 vaccination campaign in NYC averted an average of 252,737 cases (95% CrI: 217,503 - 289,582), 44,211 hospitalizations (95% CrI: 39,102 - 49,084) and 8,306 deaths (95% CrI: 7,129 - 9,482), corresponding to a reduction of 29.7% cases, 50.9% hospitalizations and 48.1% deaths expected during the evaluation period (Figure 1).

**Figure 1.**
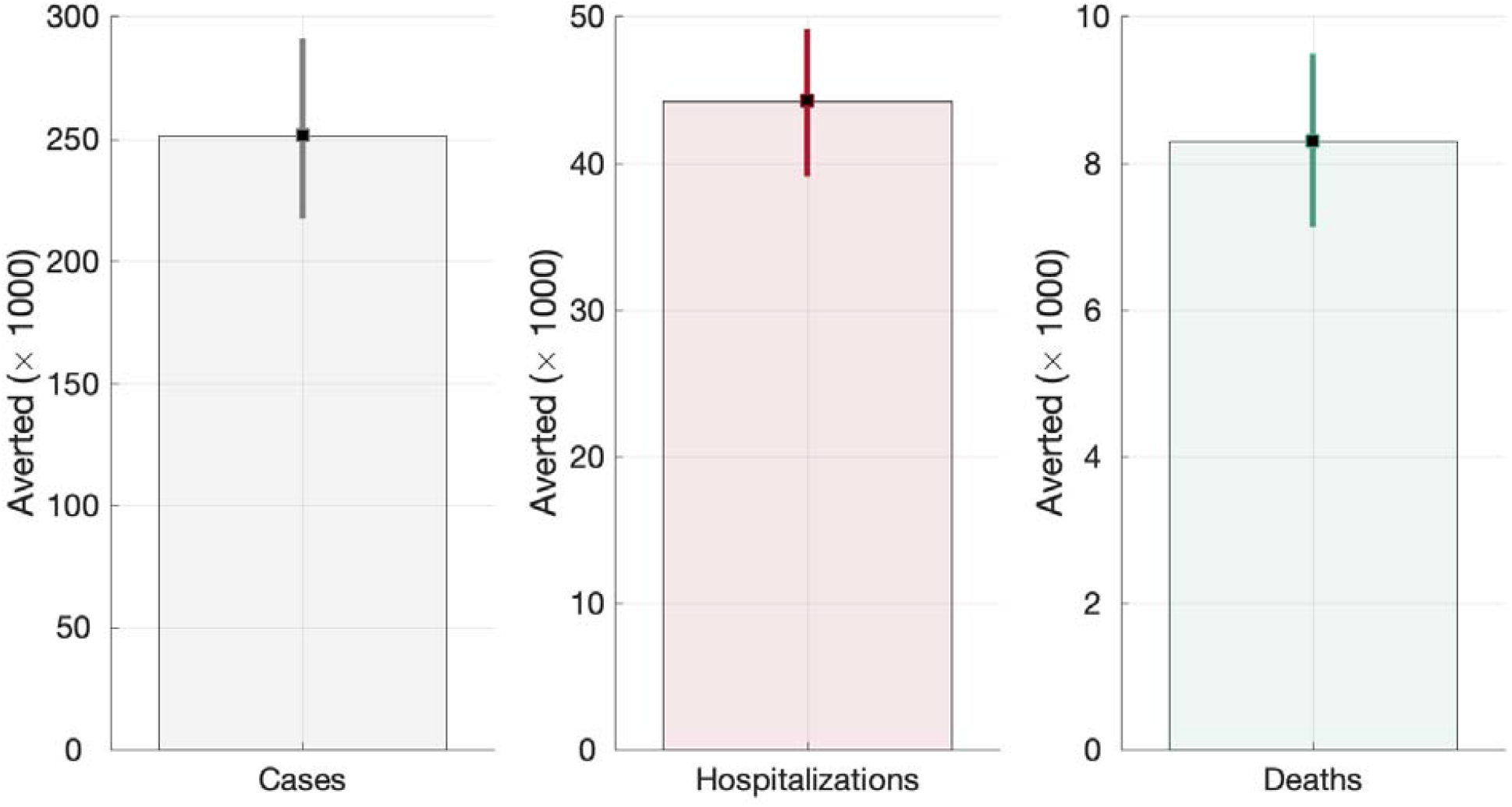
Averted cases, hospitalizations and deaths with vaccination compared to no vaccination since the start of vaccine rollout through July 1, 2021.

Hospitalizations and deaths were reduced to a higher degree compared with incidence, likely due to higher efficacy of the vaccines against symptomatic and severe disease than infection and transmission. Furthermore, vaccine rollout prioritized individuals at higher risk of severe illness and deaths from COVID-19. Notably, the swift vaccine rollout suppressed another wave of COVID-19 that would have led to sustained increase in cases, hospitalizations and deaths during spring triggered by highly transmissible variants (Figures 2-4). In particular, without vaccination, Alpha variant would have caused an average of more than 950 hospitalizations and 215 deaths per day at the peak of the resurgence in April 2021, which is twice the previous peak of hospitalizations and deaths in January, 2021. Continued vaccination also curbed elevated hospitalizations and deaths expected despite the increasing dominance of the Delta variant starting from May.

**Figure 2.**
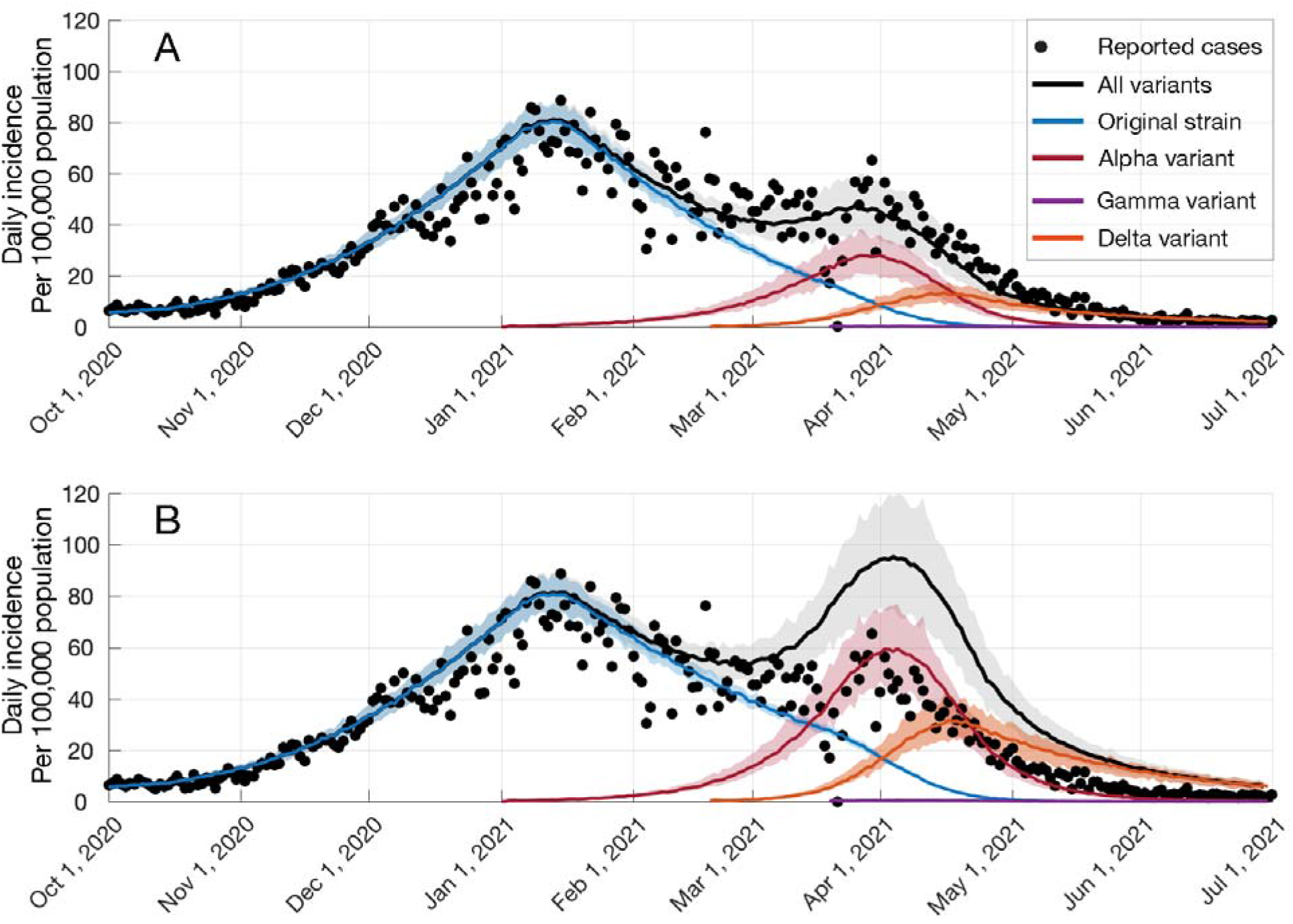
Impact of COVID-19 vaccination progress. Projected daily incidence per 100,000 population (A) with vaccination; (B) without vaccination

**Figure 3.**
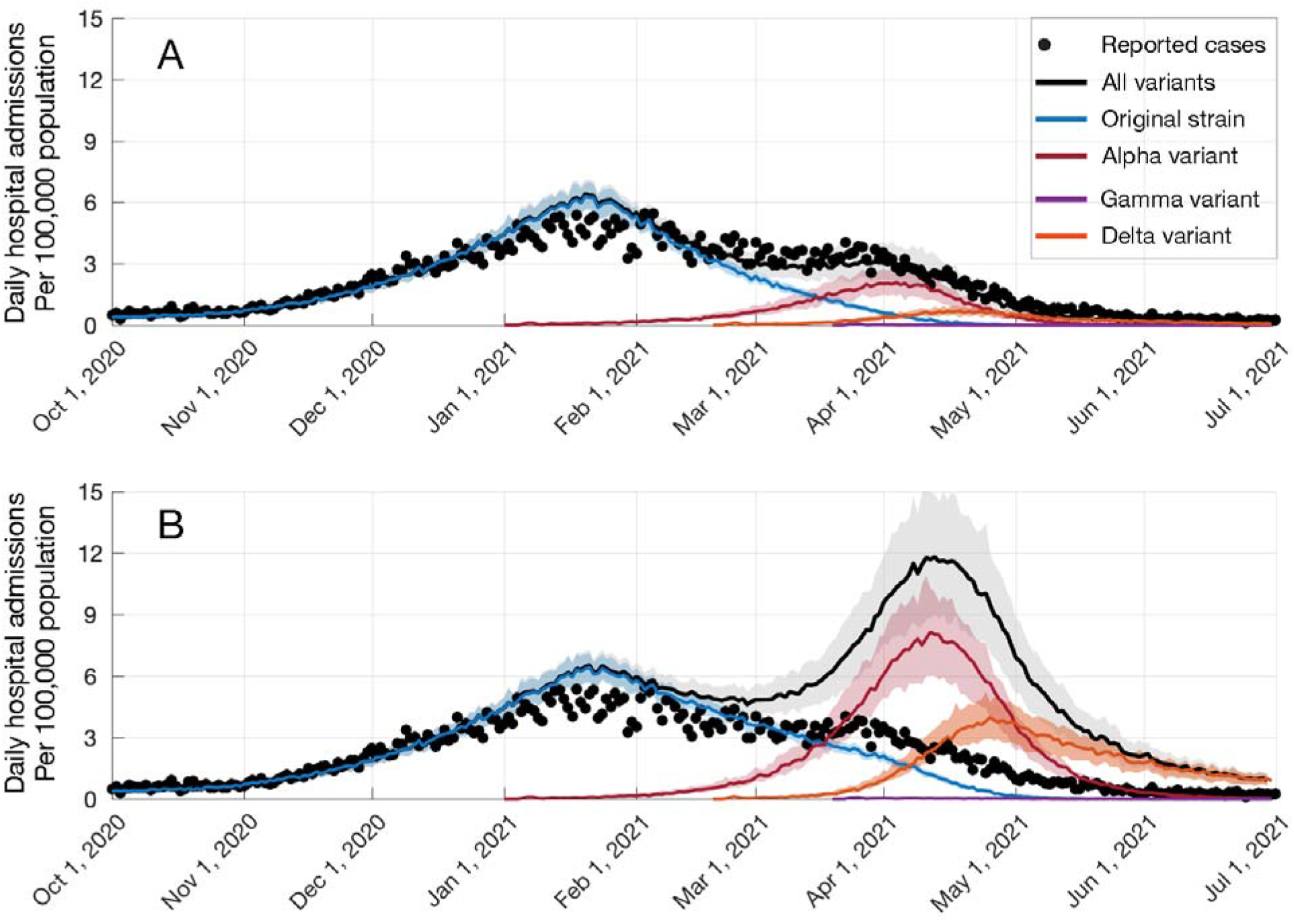
Impact of COVID-19 vaccination progress. Projected daily hospital admissions per 100,000 population (A) with vaccination; (B) without vaccination

**Figure 4.**
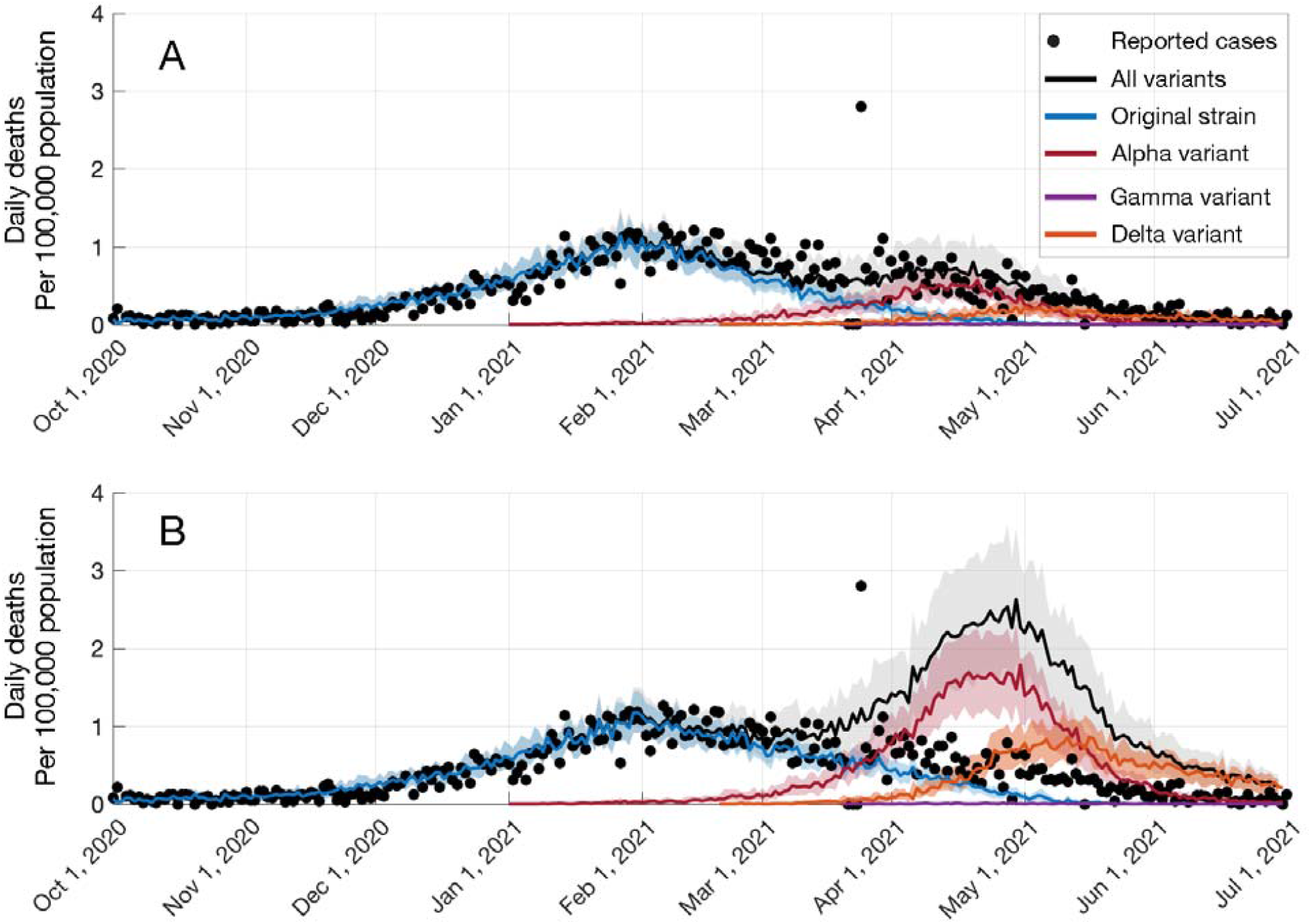
Impact of COVID-19 vaccination progress. Projected daily deaths per 100,000 population (A) with vaccination; (B) without vaccination

## Discussion

Our study underscores that the swift vaccine rollout in NYC played a pivotal role in reducing COVID-19 burden as well as suppressing potential outbreaks due to more transmissible emerging variants. Nevertheless, highly transmissible variants, such as Delta, are currently sweeping across the city with over a three-fold increase in the average incidence from June to July. This highlights the continued need to expand vaccination as well as focusing on underserved neighbourhoods and communities with a low-vaccination coverage to achieve outbreak control and maximize protection against future variants.

## Data Availability

All data is publicly available

## Acknowledgements

Alexandra Ternier, Vassiliki Papadouka, Iris Cheng, Mohammed Almashhadani, Ariel Charney, Charles Ko, and colleagues across the Bureau of Immunization and Integrated Data Team for processing, preparing, and analyzing vaccination data. Sharon Greene, Corinne Thompson and colleagues in Surveillance and Epidemiology and the Public Health Laboratory for processing, preparing, and analyzing data on COVID-19 cases, hospitalizations, and deaths. Scott Harper and Jennifer Rosen for scientific expertise.

