## Appendix for "Lives saved and hospitalizations averted by COVID-19 vaccination in New York City"

DRAFT

**Model structure**
We extended our previous agent-based model of COVID-19 transmission and vaccination [1,2] to include B.1.1.7 (Alpha), P.1 (Gamma) and B.1.617.2 (Delta) variants of SARS-CoV-2 with different transmissibilities in addition to the original strain. The model implemented the natural history of disease with epidemiological classes for susceptible; latently infected (not yet infectious); asymptomatic (and infectious); pre-symptomatic (and infectious); symptomatic (and infectious) with either mild or severe illness; recovered; and dead. The population was stratified into different age groups and incorporated age-specific risk of hospitalizations and deaths, contact patterns, and a two-dose vaccination rollout. Daily contacts between individuals were sampled from a negative-binomial distribution parameterized (Table S1) using empirical data on pre-pandemic and pandemic-era interactions .[3,4]

**Table S1.** Mixing patterns and the daily number of contacts derived from empirical observations .[3,4] Daily numbers of contacts were sampled from negative binomial distributions for different scenarios.

| Age group | Proportion of contacts between age groups | | | | | No. of daily contacts without self-isolation Mean (SD) | No. of daily contacts for self-isolated individuals Mean (SD) |
|---|---|---|---|---|---|---|---|
| | 0-4 | 5-19 | 20-49 | 50-65 | 65+ | | |
| 0-4 | 0.2287 | 0.1839 | 0.4219 | 0.1116 | 0.0539 | 10.21 (7.65) | 2.86 (2.14) |
| 5-19 | 0.0276 | 0.5964 | 0.2878 | 0.0591 | 0.0291 | 16.793 (11.7201) | 4.70 (3.28) |
| 20-49 | 0.0376 | 0.1454 | 0.6253 | 0.1423 | 0.0494 | 13.795 (10.5045) | 3.86 (2.95) |

| | | | | | | | |
|---|---|---|---|---|---|---|---|
| 50-65 | 0.0242 | 0.1094 | 0.4867 | 0.2723 | 0.1074 | 11.2669 (9.5935) | 3.15 (2.66) |
| 65+ | 0.0207 | 0.1083 | 0.4071 | 0.2193 | 0.2446 | 8.0027 (6.9638) | 2.24 (1.95) |

**Transmissibility**
Risk of infection for susceptible individuals depended probabilistically on their interaction with infectious individuals in the pre-symptomatic, symptomatic, or asymptomatic stages of infection. The transmission probability of the original strain of SARS-CoV-2 was calibrated by fitting the model to case incidence data per 100,000 population in the entire US from October 1, 2020, to July 1, 2021.[5] We chose October 1 as the starting point for our calibration and simulations because it was a time of a relatively low incidence preceding the fall/winter wave in the US. The calibration (in the presence of only the original strain of SARS-CoV-2) resulted in a transmission probability of 0.109. This transmission probability corresponds to an effective reproduction number of 1.17 in early October 2020 [6], which accounted for the effect of non-pharmaceutical interventions (NPIs) in simulated scenarios. We then introduced the Alpha variant on January 3, 2021 with a 50% higher transmissibility compared to the original strain. [7–9] We introduced the Gamma variant on March 20, 2021 and the Delta variant (B.1.617.2) in the model on February 20, 2021, as reported by the NYC Department of Health and Mental Hygiene.[10] The transmissibility of this variant was set as 30% higher relative to the Alpha variant. [11]

**Disease dynamics**
We parameterized the infectivity of asymptomatic, mild symptomatic, and severe symptomatic individuals to be 26%, 44%, and 89% relative to the pre-symptomatic stage.[12–14] We assumed that these relative infectivities remained the same for all variants in the model. The incubation period was sampled from a log-normal distribution with a mean of 5.2 days,[15] and parameters of 1.434 (shape) and 0.661 (scale). An age-dependent proportion of infected individuals progressed to a pre-symptomatic stage with a mean duration of 2.3 days, sampled from a Gamma distribution with parameters of 1.058 (shape) and 2.17 (scale). [13,16] Pre-symptomatic cases developed symptomatic disease with a mean duration of 3.2 days, which was also sampled from a Gamma distribution with parameters of 2.77 (shape) and 1.1563 (scale).[17,18] The remaining proportion of infected individuals experienced asymptomatic infection until recovery, with a mean infectious period of 5 days sampled from a Gamma distribution with parameters of 5 (shape) and 1 (scale). [17,18]

Recent studies indicate that antibodies from prior infection with other variants of SARS-CoV-2 may have reduced neutralizing activity against Gamma and Delta.[19–22] We therefore assumed that both the Gamma and Delta variants evade naturally acquired immunity by an average of 21% (95% CI: 11-36%).[23,24] This evasion rate was implemented as a reduction of immune protection for individuals recovered from the original strain or the Alpha variant, corresponding to an average transmission probability of $0.21\times0.109 = 0.0229$. We further assumed that recovery from infection due to the Gamma or Delta variant provides protection against all variants in the model, preventing reinfection for at least one year.

**Infection outcomes**
We assumed that asymptomatic and mild symptomatic cases recover from infection without hospitalization. A proportion of those with severe disease were hospitalized within 2-5 days of symptom onset [25,26] and were therefore removed from the transmission chain. We also assumed that all symptomatic cases who were not hospitalized self-isolated within 24 hours of symptom

onset, and reduced their number of daily contacts by an additional 72% (Table S1). Intensive care unit (ICU) and non-ICU hospitalization rates were parameterized (Table S2) by clinical and epidemiological data stratified by age and comorbidities.[27–29] Infection with Alpha variant was associated with 64% higher risk of death,[8,9] and infections with Gamma or Delta variants were assigned the case fatality of the original strain.

**Table S2.** Model parameters associated with hospitalization of severe cases.

| | | | |
|---|---|---|---|
| Proportion of severe cases hospitalized with one or more comorbidities | | 100% | [27–29] |
| | Non-ICU | 60.4% | |
| | ICU | 39.6% | |
| Proportion of severe cases hospitalized without any comorbidities | | 10.8% | [27–29] |
| | Non-ICU | 75% | |
| | ICU | 25% | |
| Length of non-ICU stay (days) | | Gamma(shape: 4.5, scale: 2.75) | Derived from [30,31] |
| Length of ICU stay (days) | | Gamma(shape: 4.5, scale: 2.75) + 2 | Derived from [30,31] |

### Vaccination

We implemented a two doses vaccination campaign following the reported daily distribution of vaccines per age group in NYC [32]. We assumed that comorbid individuals had priority within the age groups. We specified Pfizer-BioNTech vaccines with an interval of 21 days between the first and second doses. [33] This interval was 28 days for Moderna vaccines. [34] We parameterized the model with published estimates of vaccine efficacy following each dose of Pfizer-BioNTech and Moderna vaccines against infection, symptomatic disease, and severe disease caused by the original strain.[1,35] These efficacies, reported in Table S3, were implemented in the model as a reduction of transmission probability (for efficacy against infection), reduction in probability of developing symptomatic disease, and reduction of severe illness if symptomatic disease occurred. For efficacy of Pfizer-BioNTech vaccines against infection and severe disease, we used recent estimates to inform the model.[36]

Table S3. Estimated vaccine efficacies (%) from published studies.

| **Vaccine efficacy** Pfizer-BioNTech | **Weeks after the first dose** | | **Weeks after the second dose** | | **Reference** |
|---|---|---|---|---|---|
| Original strain | 1-2 | 3 | 1-2 | >2 | [34,37–40] |
| Infection | None | 46 (40, 51) | 60 (53, 66) | 92 (88, 95) | |

| | | | | | |
|---|---|---|---|---|---|
| Symptomatic disease | None | 57 (50, 63) | 66 (57, 73) | 94 (87, 98) | |
| Severe disease | None | 62 (39, 80) | 80 (59, 94) | 92 (75, 100) | |
| Alpha variant | 1-2 | 3 | 1-2 | >2 | |
| Infection | None | 29.5 (22.9, 35.5) | 60 (53, 66) | 89.5 (85.9, 92.3) | 36,41 |
| Symptomatic disease | None | 53.6 (50, 63) | 62 (57, 73) | 93.4 (90.4, 95.5) | |
| Severe disease | None | 54.1 (26.1, 71.9) | 80 (59, 94) | 94 (87, 98) | |
| Beta/Gamma variant * | 1-2 | 3 | 1-2 | >2 | |
| Infection | None | 36.8 (32, 40.8)** | 48 (42.4, 52.8)** | 73.6 (70.4, 76)** | 36,41 |
| Symptomatic disease | None | 33.2 (8.3, 51.4) | 66 (57, 73) | 94 (87, 98) | |
| Severe disease | None | 34 (0, 50) | 68 (64, 75) | 97.4 (92.2, 99.5) | |
| Delta variant | 1-2 | 3 | 1-2 | >2 | |
| Infection | None | 36.8 (32, 40.8)** | 48 (42.4, 52.8)** | 73.6 (70.4, 76)** | 41 |
| Symptomatic disease | None | 33.2 (8.3, 51.4) | 62 (57, 73) | 93.4 (90.4, 95.5) | |
| Severe disease | None | 34 (0, 50) | 68 (64, 75) | 97.4 (92.2, 99.5) | |

* Vaccine efficacy against the Gamma variant was assumed to be the same as those reported for Beta.
** 20% of escaping compared to original strain.

**Model implementation**
Assuming 10% pre-existing immunity generated by the original strain prior to October 2020, [42,43] we simulated the model with a population of 100,000 individuals from September 1, 2020 to December 1, 2021. To incorporate the age distribution of pre-existing immunity in the population, we ran the model with only the original strain in the absence of vaccination and determined the infection rates in different age groups when the overall attack rate reached 10%. The distribution of this immunity was used to parameterize the initial population at the start of simulations. Vaccination was initiated on December 12, and rolled out as a two-dose strategy.

On April 2, the guidelines by the US Centers for Disease Control and Prevention indicated a minimal risk for fully vaccinated individuals to travel and engage in certain social activities while taking COVID-19 precautions.[44] We therefore allowed vaccinated individuals to return to normal pre-pandemic behaviour 14 days after the second dose of vaccine from April 3, 2021. The model was implemented in Julia, which is an open-source, high-performance, dynamic programming language that allows rapid analysis of computationally intensive problems, such as agent-based modelling.

13 Moghadas SM, Fitzpatrick MC, Sah P, *et al.* The implications of silent transmission for the

control of COVID-19 outbreaks. *Proc Natl Acad Sci U S A* 2020; **117**: 17513–5.

14 Sayampanathan AA, Heng CS, Pin PH, Pang J, Leong TY, Lee VJ. Infectivity of asymptomatic versus symptomatic COVID-19. *Lancet* 2021; **397**: 93–4.

15 Li Q, Guan X, Wu P, *et al.* Early Transmission Dynamics in Wuhan, China, of Novel Coronavirus-Infected Pneumonia. *N Engl J Med* 2020; **382**: 1199–207.

16 He X, Lau EHY, Wu P, *et al.* Temporal dynamics in viral shedding and transmissibility of COVID-19. *Nat Med* 2020; **26**: 672–5.

17 Li R, Pei S, Chen B, *et al.* Substantial undocumented infection facilitates the rapid dissemination of novel coronavirus (SARS-CoV-2). *Science* 2020; **368**: 489–93.

18 Gatto M, Bertuzzo E, Mari L, *et al.* Spread and dynamics of the COVID-19 epidemic in Italy: Effects of emergency containment measures. *Proc Natl Acad Sci U S A* 2020; **117**: 10484–91.

19 Zhou D, Dejnirattisai W, Supasa P, *et al.* Evidence of escape of SARS-CoV-2 variant B.1.351 from natural and vaccine-induced sera. *Cell* 2021; **184**: 2348–61.e6.

20 Planas D, Bruel T, Grzelak L, *et al.* Sensitivity of infectious SARS-CoV-2 B.1.1.7 and B.1.351 variants to neutralizing antibodies. *Nat Med* 2021; published online March 26. DOI:10.1038/s41591-021-01318-5.

21 Vaidyanathan G. Coronavirus variants are spreading in India - what scientists know so far. *Nature* 2021; **593**: 321–2.

22 Liu C, Ginn HM, Dejnirattisai W, *et al.* Reduced neutralization of SARS-CoV-2 B.1.617 by vaccine and convalescent serum. *Cell* 2021; published online June 17. DOI:10.1016/j.cell.2021.06.020.

23 Centre for Mathematical Modelling of Infectious Diseases. Estimates of severity and transmissibility of novel SARS-CoV-2 variant 501Y.V2 in South Africa. Estimates of severity and transmissibility of novel SARS-CoV-2 variant 501Y.V2 in South Africa. 2021; published online Jan 11. https://cmmid.github.io/topics/covid19/sa-novel-variant.html (accessed May 2, 2021).

24 Jangra S, Ye C, Rathnasinghe R, *et al.* SARS-CoV-2 spike E484K mutation reduces antibody neutralisation. *Lancet Microbe* 2021; **2**: e283–4.

25 Shoukat A, Wells CR, Langley JM, Singer BH, Galvani AP, Moghadas SM. Projecting demand for critical care beds during COVID-19 outbreaks in Canada. *CMAJ* 2020; **192**: E489–96.

26 Moghadas SM, Shoukat A, Fitzpatrick MC, *et al.* Projecting hospital utilization during the COVID-19 outbreaks in the United States. *Proc Natl Acad Sci U S A* 2020; **117**: 9122–6.

27 Garg S, Kim L, Whitaker M, *et al.* Hospitalization rates and characteristics of patients hospitalized with laboratory-confirmed Coronavirus disease 2019 - COVID-NET, 14 states, March 1-30, 2020. *MMWR Morb Mortal Wkly Rep* 2020; **69**: 458–64.

28 Team CC-19 R, CDC COVID-19 Response Team, Chow N, *et al.* Preliminary Estimates of

the Prevalence of Selected Underlying Health Conditions Among Patients with Coronavirus Disease 2019 — United States, February 12–March 28, 2020. MMWR. Morbidity and Mortality Weekly Report. 2020; **69**: 382–6.

29 Nyberg T, Twohig KA, Harris RJ, *et al.* Risk of hospital admission for patients with SARS-CoV-2 variant B.1.1.7: cohort analysis. *BMJ* 2021; **373**: n1412.

30 Guan W-J, Ni Z-Y, Hu Y, *et al.* Clinical Characteristics of Coronavirus Disease 2019 in China. *N Engl J Med* 2020; **382**: 1708–20.

31 Sanche S, Lin YT, Xu C, Romero-Severson E, Hengartner N, Ke R. High Contagiousness and Rapid Spread of Severe Acute Respiratory Syndrome Coronavirus 2. *Emerg Infect Dis* 2020; **26**: 1470–7.

32 coronavirus-data. Github https://github.com/nychealth/coronavirus-data (accessed July 13, 2021).

33 Anderson EJ, Rouphael NG, Widge AT, *et al.* Safety and Immunogenicity of SARS-CoV-2 mRNA-1273 Vaccine in Older Adults. *N Engl J Med* 2020; **383**: 2427–38.

34 Polack FP, Thomas SJ, Kitchin N, *et al.* Safety and Efficacy of the BNT162b2 mRNA Covid-19 Vaccine. *N Engl J Med* 2020; **383**: 2603–15.

35 Moghadas SM, Vilches TN, Zhang K, *et al.* Evaluation of COVID-19 vaccination strategies with a delayed second dose. *PLoS Biol* 2021; **19**: e3001211.

36 Abu-Raddad LJ, Chemaitelly H, Butt AA, National Study Group for COVID-19 Vaccination. Effectiveness of the BNT162b2 Covid-19 Vaccine against the B.1.1.7 and B.1.351 Variants. *N Engl J Med* 2021; published online May 5. DOI:10.1056/NEJMc2104974.

37 Dagan N, Barda N, Kepten E, *et al.* BNT162b2 mRNA Covid-19 Vaccine in a Nationwide Mass Vaccination Setting. *N Engl J Med* 2021; **384**: 1412–23.

38 U.S. Food and Drug Administration. Vaccines and related biological products advisory committee December 10, 2020 meeting briefing document. U.S. Food and Drug Administration, 2020 https://www.fda.gov/media/144246/ (accessed July 2, 2021).

39 Lipsitch M, Kahn R. Interpreting vaccine efficacy trial results for infection and transmission. *Vaccine* 2021; published online June 12. DOI:10.1016/j.vaccine.2021.06.011.

40 Chodick G, Tene L, Patalon T, *et al.* Assessment of Effectiveness of 1 Dose of BNT162b2 Vaccine for SARS-CoV-2 Infection 13 to 24 Days After Immunization. *JAMA Netw Open* 2021; **4**: e2115985.

41 Vizient, Inc. COVID-19 vaccine candidates. 2021 https://www.vizientinc.com/-/media/documents/sitecorepublishingdocuments/public/covid19_sidebyside_vaccinecompare.pdf.

42 Bajema KL, Wiegand RE, Cuffe K, *et al.* Estimated SARS-CoV-2 Seroprevalence in the US as of September 2020. *JAMA Intern Med* 2021; **181**: 450–60.

43 Arora R, Yan T. SeroTracker. SeroTracker. 2021; published online May 2. https://serotracker.com (accessed May 2, 2021).

44 CDC. Interim public health recommendations for fully vaccinated people. 2021; published online April 30. https://www.cdc.gov/coronavirus/2019-ncov/vaccines/fully-vaccinated-guidance.html (accessed May 2, 2021).
